# A prospective pilot study demonstrating non-invasive calibration-free glucose measurement

**DOI:** 10.1101/2024.08.17.24312052

**Authors:** Martina Rothenbühler, Aritz Lizoain, Fabien Rebeaud, Adler Perotte, Marc Stoffel, J. Hans DeVries

## Abstract

Glucose is an essential molecule in energy metabolism. Dysregulated glucose metabolism, the defining feature of diabetes, requires active monitoring to prevent significant morbidity and mortality. Current technologies for intermittent and continuous glucose measurement are invasive. Non-invasive glucose measurement would eliminate this barrier towards making glucose monitoring more accessible, extending the benefits from people living with diabetes to prediabetes and the healthy. We developed and investigated a spectroscopy-based system for measuring glucose non-invasively and without per-person calibration. Using data from a study including adults with insulin-treated diabetes, we constructed a computational model from a development cohort of 15 subjects and found a mean absolute relative difference of 14.5% in an independent validation cohort of five subjects. The correlation between the average model sensitivity by wavelength and the spectrum of glucose was 0.45 (p<0.001). Our findings suggest that spectroscopy-based non-invasive measurement of glucose without invasive calibration is possible.

## Main

Glucose is a tightly regulated energy source for human cells, fueling many cellular processes and metabolic functions. Blood glucose concentration affects and is affected by many factors including satiety, exercise, sleep, hormones, and mood^1^. Daily blood glucose fluctuations are a normal response to dietary intake and energy expenditure, but an excess caloric intake particularly rich in sugars and starches combined with a sedentary lifestyle are proven risk factors for the development of dysglycemia and, eventually, diabetes mellitus (DM)^2^. DM is a notable health concern worldwide, currently affecting approximately half a billion people, projected to reach 700 million by 2045^3^ with significant morbidity and mortality largely caused by micro- and macrovascular complications.

The real-time tracking of glucose via continuous glucose monitors (CGMs) has improved the management of DM, particularly in type 1 and insulin-treated 2 DM, where reductions of HbA1c, increases in time spent in the target glucose range, and reductions in the number of hypoglycemic episodes have been demonstrated^4–7^. More recent data shows that benefits are also seen in non-insulin-treated type 2 DM^7,8^, while the value of CGM in prediabetes^9^, and in people without dysglycemia^10,11^ is an area of active research. Supporting this trend, the FDA has recently cleared the use of CGMs for people without diabetes^12^.

Today’s CGMs are invasive, relying on a needle-based applicator for the insertion of a microneedle filament, which uses an electrochemical approach to quantify the glucose concentration in the interstitial fluid. Since their market entry, CGM devices have steadily increased both their accuracy and lifetime, reduced their footprint, and expanded their connectivity to phones, smartwatches, and pumps. Although invasive CGMs have greatly improved the quality of care for people living with insulin-treated DM, non-invasive CGM technology can eliminate the discomfort and risks associated with invasive monitoring, thus eliminating a significant barrier to adoption^13^. However, in order to truly eliminate this barrier, a non-invasive monitoring approach should also not require per-person invasive calibration to continuously measure glucose accurately. Many technologies and approaches have been proposed and pursued, but a non-invasive method for glucose measurement without per-person calibration has been elusive^14^.

We here report the first results of a single-center, multiple sequential-cohort study designed to assess the ability of a novel non-invasive investigational device to measure glucose without per-person calibration. The investigational device makes optical measurements based on the principle of Raman spectroscopy. Reference venous plasma glucose measurements and spectral data from the investigational device were collected for up to 6 hours on adults living with type 1 DM who underwent a carbohydrate-rich meal challenge with delayed insulin bolus administration (see Methods). Twenty participants were enrolled and completed the study procedures according to protocol. The first 15 subjects were allocated to the development cohort and the following five participants were allocated to the validation cohort. There was a strict separation of investigators involved in model development (FR, AP) and validation (MR). The reference plasma glucose values of the validation cohort were generated and retained by the validation investigators to ensure an independent external assessment of the performance of the investigational device. This pre-defined, mutually exclusive data split also eliminated the possibility of per-person calibration.

The study population consisted of ten women and ten men, median age was 41 years (IQR 31 to 45), median BMI was 25.8 kg/m^2^ (IQR 22.9 to 28.6) and median HbA1c was 7.3% (IQR 6.7 to 7.7). All participants had type 1 diabetes since more than one year and skin type II (25%), III (40%) or IV (35%) according to the Fitzpatrick scale^15^. Among all participants, time from start of intervention to hyperglycaemia ranged from 5 to 75 minutes and median time in hyperglycaemia was 260 minutes (IQR 197.5 to 274.3). Median plasma glucose values were 6.5 mmol/l (IQR 5.8 to 7.5) at the start of the procedure and similar (6.6 mmol/l; IQR 4.7 to 10) at discharge. Median plasma glucose during hyperglycaemia was 14.2 mmol/l (IQR 13.3 to 15.5). The lowest measured plasma glucose value among all interventions was 3.2 mmol/l, and the highest 16.7 mmol/l. We categorized the rate of change (RoC) into low (less than 0.06 mmol/l/min), moderate (between 0.06 and 0.11 mmol/l/min), high (between 0.11 and 0.17 mmol/l/min) and very high (above 0.17 mmol/l/min)^16^. Distributions across the different RoC categories were comparable in the two cohorts. In both groups, 16% of the measured datapoints were during very high RoC, approximately 15% in high, 31% in moderate and 37% during low RoC.

The mean absolute relative difference (MARD) in the validation cohort was 14.5%, ranging from 11.1% to 22.03% at the individual level. The MARD in hyperglycemia (> 10 mmol/l) was 12.4% (range at the individual level: 7.0% to 19.9%) and the MARD in euglycemia (3.9 to 10.0 mmol/l) was 25.9% (range at the individual level: 12.1% to 40.8%). MARD during low RoC was 14.9%, 11.6% during moderate, 11.2% during high and 19.6% during very high RoC. The Parkes error grid of the entire validation set is shown in Figure 2^17^. 80% of matched pairs were in zone A, 18% in zone B and 2% in zone C, while none in zones D and E. Four out of the five participants had estimated glucose values that were in zones A or B only. The individual proportion of data points in zone A ranged from 70.2% to 90.7% (see suppl. Figure). When considering the RoC, 81% of the observations with low, 85% with moderate, 87% with high and 67% with very high RoC were in zone A. All data points in zone C were measured during very high RoC.

**Figure 1.**
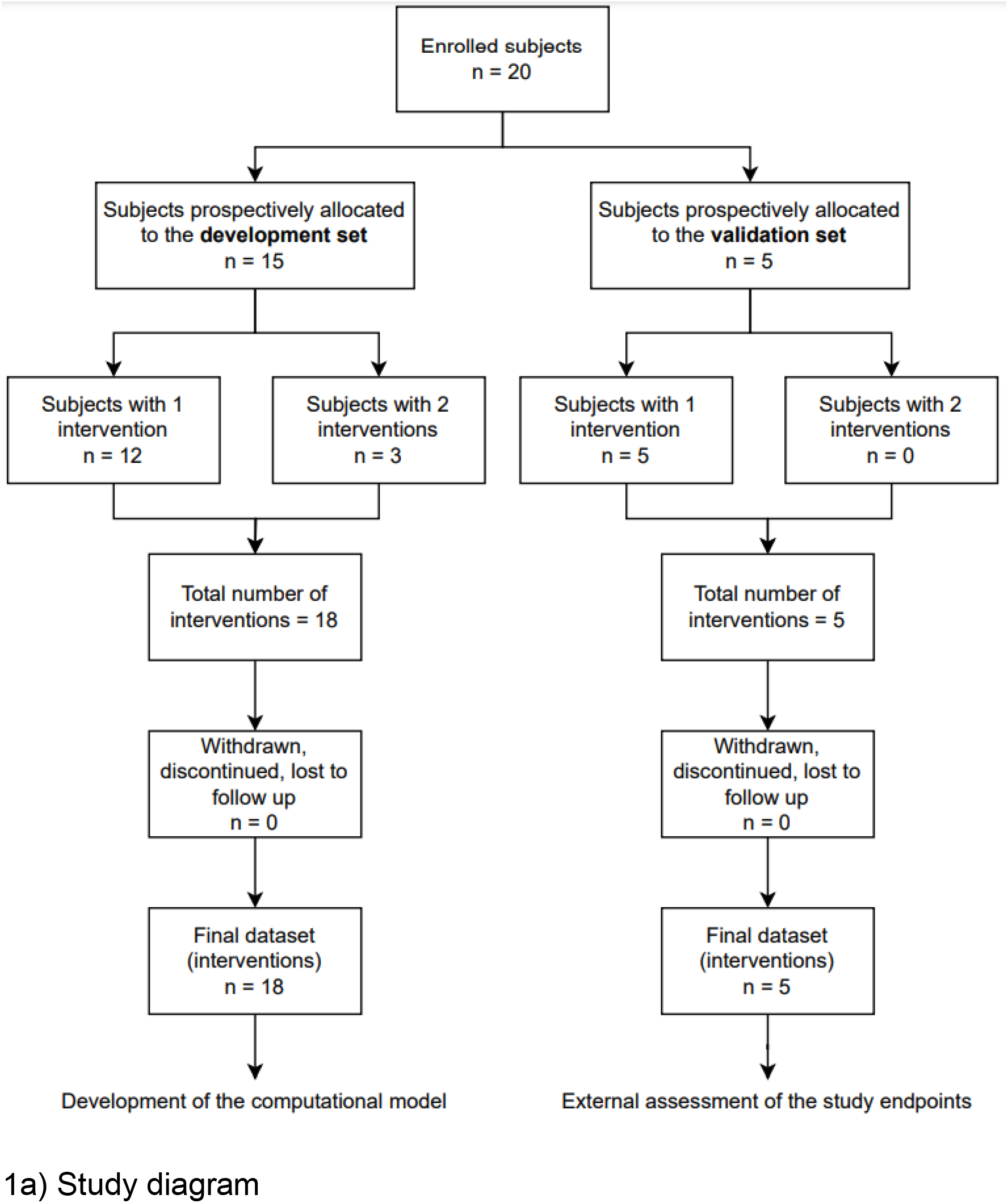
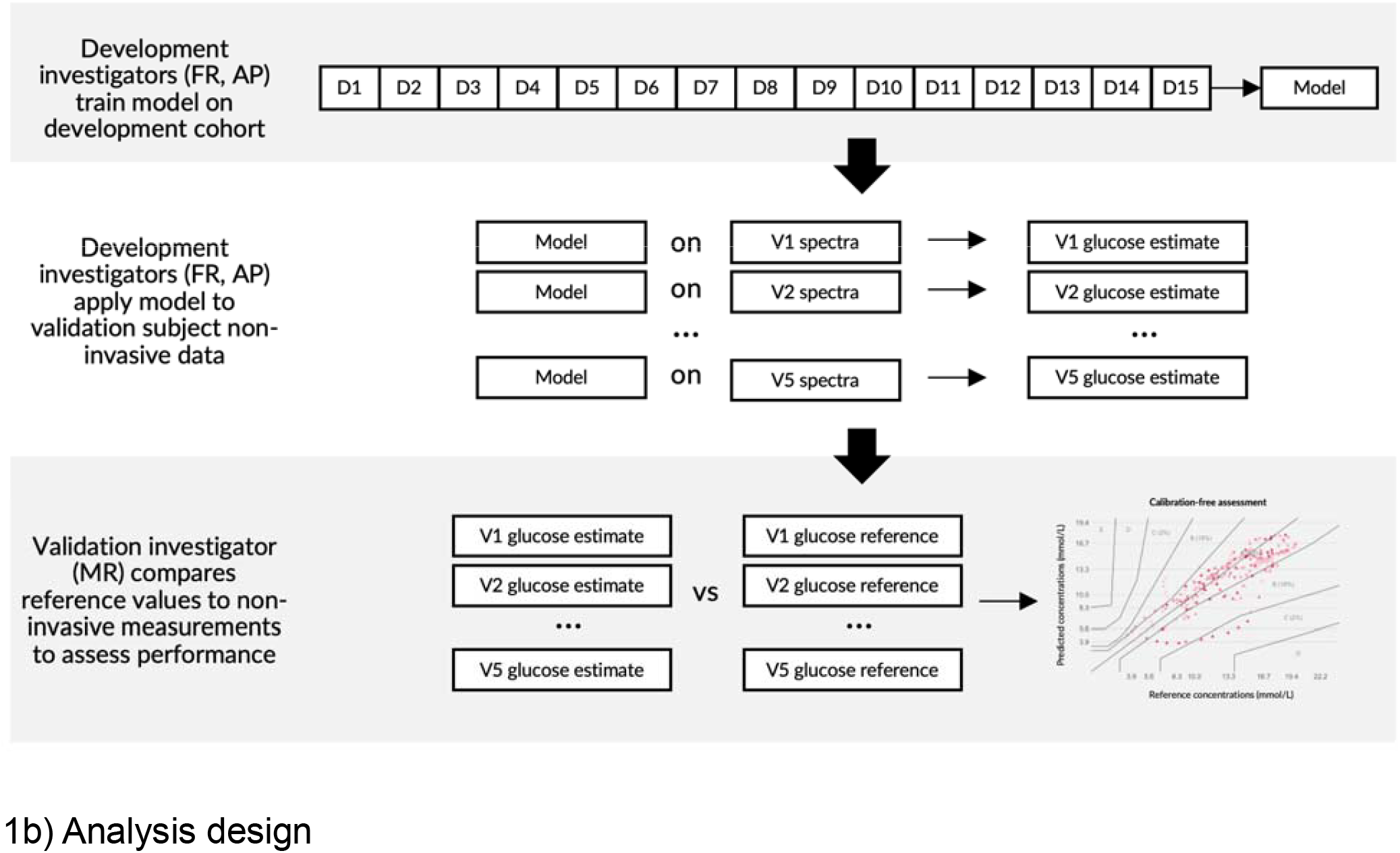
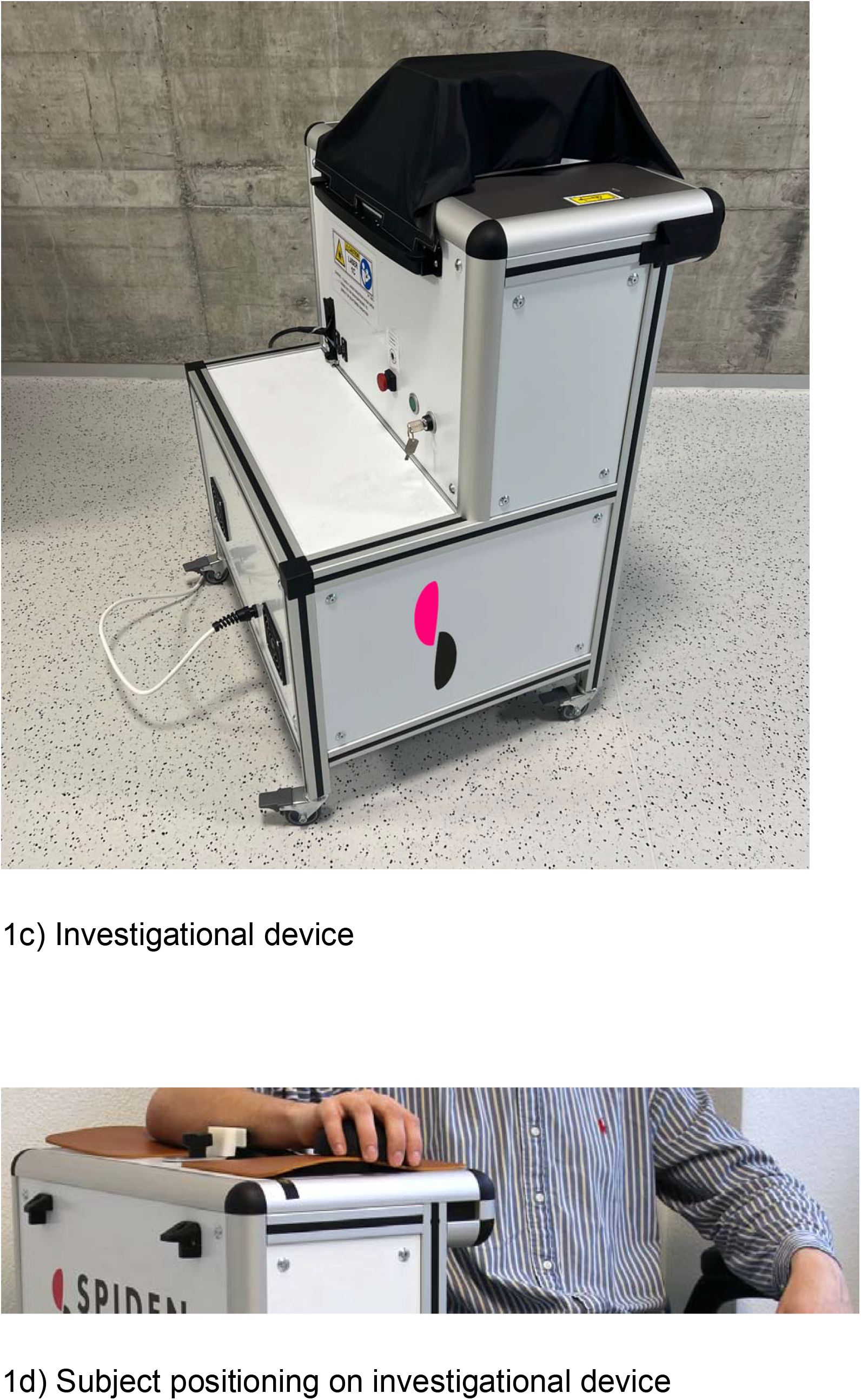

**Figure 2.**
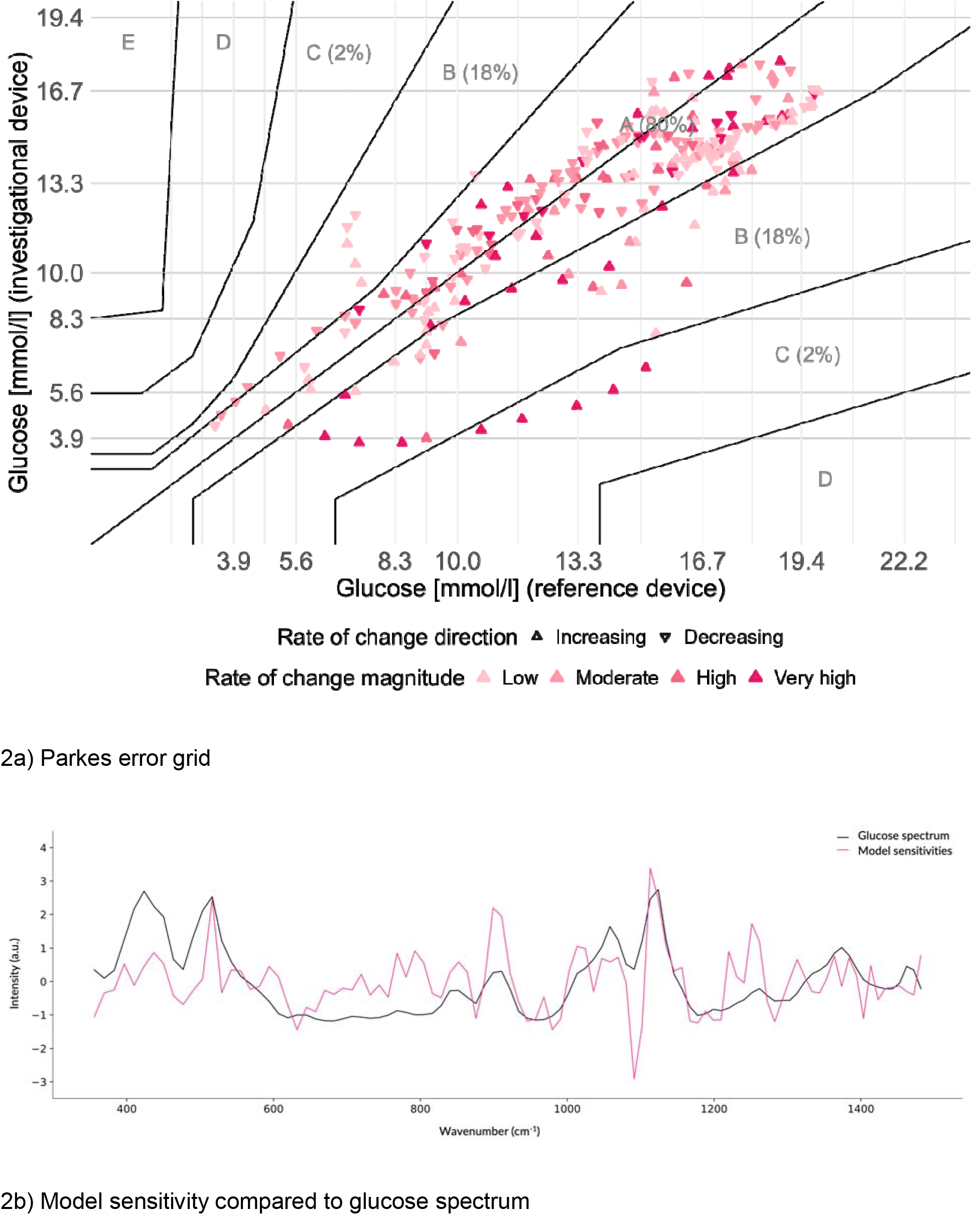

Due to the nature of these non-invasive measurements, there is a question of whether the estimates of glucose are based on a correlate of glucose or based on a measurement of glucose itself. To investigate this, we evaluated our model’s average glucose estimate sensitivity to changes in Raman spectra (i.e., the gradient of the independent variable of glucose with respect to the dependent variables of Raman spectrum intensities) and compared this to the glucose spectrum as evaluated from a series of aqueous glucose solutions (Figure 2). To provide robust estimates of glucose, the model must not only be sensitive to glucose, but also insensitive to other Raman signals unrelated to glucose estimation. Due to this dual role, it is not expected that the correlation coefficient is close to one, but ideally has a moderate value instead. We found a Pearson correlation coefficient of 0.45 (p<0.001).

In summary, 98% of the matched pairs between estimated and reference plasma glucose values were in zone A and B of the Parkes error grid, which represent the zones where errors are expected to have little or no effect on clinical outcome. The accuracy was relatively consistent across different RoC and ranged from an MARD of 11.2% during high and 19.6% during very high RoC.

One limitation of our study is the modest sample size of the validation cohort and the lack of subjects with darker skin (types V and VI on the Fitzpatrick scale). In addition, there is only limited data in hypoglycemia. Future work will aim at replicating these results in a more diverse population, including the assessment of the system’s performance in healthy and prediabetes populations.

The findings of our study suggest that measuring glucose noninvasively without invasive calibration is possible and confirm that optical systems combined with advanced computational models are highly promising. Overall, the performance is comparable to first generation CGMs^18^, but unlike those systems, this device does not require per-person calibration. Additionally, the accuracy of the investigational device was further challenged, and its performance confirmed by the study procedure, which consisted of rapid and large glucose excursions in a population of people living with profound defects in their glucose metabolism, making glucose monitoring especially challenging. The convergence of recent developments in the fields of photonics, electronics, and computational sciences, especially regarding miniaturization, open new opportunities to bring continuous, noninvasive glucose sensing to the wrist eventually. The results presented here lay down the foundations toward the development of such a technology.

## Data Availability

All validation data produced in the present work are contained in the manuscript

