## Supplementary material for "A prospective pilot study demonstrating non-invasive calibration-free glucose measurement": Online methods

This is an exploratory, prospective, single-centre, multiple-cohorts study conducted at Profil (Neuss, Germany) under consideration of the Declaration of Helsinki and in compliance with the Guideline for Good Clinical Practice and the applicable national regulations and provisions. The study protocol was approved by the responsible Ethics Committee and exempted from approval by the German Federal Institute for Drugs and Medical Devices (BfArM). The study aims to assess the accuracy in the normo- to hyperglycemic range of a novel investigational device for the non-invasive measurement of glucose in adults living with Type 1 or Type 2 diabetes (NCT06272136). The study's objectives are to assess the feasibility of the investigational device and associated computational models to detect and track glucose changes noninvasively and transcutaneously in dynamic states of glycemia, to determine the accuracy of the computational model against reference, match-paired glucose values, and to examine the safety and tolerability of the investigational device. We report here the first results issued from cohorts I and II. The first screening visit took place on February 21, 2024, and the last intervention was on June 4, 2024.

**Study devices**

The investigational device ("Clinical Demo 2.0", Liom (Spiden AG), Switzerland) is a stand-alone device with a single millimeter-size exit glass window allowing measurement on the volar surface of the wrist of human subjects. The device employs vibrational optical spectroscopy to capture the signature of the glucose molecule in the human skin in contact with the window. During the measurements, the skin is continuously exposed with a sub-millimeter near-infrared light spot at power levels below one hundred milliwatts. The device collects and captures with a spectral analyzer the near-infrared light response signals from the epidermis and dermis, which contain the instantaneous and specific signatures of the elements composing the skin tissues and liquids. The measured signals are sorted by wavenumbers, and then the corresponding amplitudes are recorded before being processed and analyzed offline with our computational procedure for glucose concentration estimation. Over the course of a visit, the glucose concentration is continuously measured at the frequency of one measure every 5 minutes to capture changes of glycemia with adequate granularity. The device is fully automatized and does not require intervention of an expert operator. The device was calibrated prior to the study but there was no intervention on the device during the whole study. The measurements were performed on the forearm of the subjects near to the wrist, where a wrist watch is typically worn, and the contact between the skin and the optical window was ensured by gravity.

Reference plasma glucose was determined using a benchtop glucose analyzer (SuperGL Compact, Dr. Müller Gerätebau GmBH, Germany) using venous whole blood, as per the device manufacturer's instructions.

**Participants**

Study participants were consenting adults living with insulin-treated type 1 or type 2 diabetes for at least 1 year and meeting all the inclusion and exclusion criteria set forth in Supplemental Table 1. Glycated hemoglobin at the screening was below or equal to 9.5%. Study participants were to have a skin tone in the Fitzpatrick scale type 1 to 4[1] and a wrist exempt from injury, infection, tattoo or any atypical skin conditions.

**Study Procedures**

Enrolled subjects were fasting for at least 8 hours prior to the intervention. An indwelling catheter for blood sampling and intravenous (IV) insulin injection was inserted into one arm, and the investigational device was fixed to the other arm. Study participants with a fasting plasma glucose ≥ 150 mg/dL received an IV insulin bolus for plasma glucose lowering. Data collection with the investigational device started 30 minutes before the baseline plasma glucose measurement. For comparison purposes, venous blood samples were collected using the indwelling catheter during the intervention. The meal test period started with plasma glucose < 150 mg/dL, and the participant received a predefined meal without insulin administration at the start of meal intake. Plasma glucose values were monitored until the level rose above 300 mg/dL. At plasma glucose ≥ 300 mg/dL, an individualized insulin bolus dose was administered subcutaneously in the abdominal area. Plasma glucose levels were monitored for 5 hours. In case plasma glucose did not rise above 300 mg/dL after meal intake, the insulin bolus was administered as soon as plasma glucose stopped rising and reached a plateau. In case plasma glucose did not drop below 150 mg/dL after insulin bolus administration, an additional insulin bolus was administered.

For safety purposes, a follow-up visit was made 5 to 8 days after the intervention day. Adverse events, including device-related events, were collected from enrollment until the end of the post-intervention follow-up.

**Data Analyses**

As this was an exploratory clinical investigation to evaluate the ability and the accuracy of the investigational device to measure glucose non-invasively, and to capture preliminary information on the investigational device to adequately plan further steps of clinical development, no sample size estimation was performed, and no formal hypothesis testing was planned.

*Baseline and procedural characteristics*

Baseline characteristics, including demographics and information on diabetes management, are presented as medians (IQR) in case of continuous variables and counts (%) in case of categorical variables. Procedural characteristics are presented as medians (IQR) and comprise durations and glucose levels in different glycemic states. In addition, we also report rates of changes, which we categorized into low (less than 0.06 mmol/l/min), moderate (between 0.06 and 0.11 mmol/l/min), high (between 0.11 and 0.17 mmol/l/min) and very high (above 0.17 mmol/l/min).

*Computational model development and validation*

Among the 20 subjects of this first study results, 15 were allocated to the training dataset, while the remaining five were allocated to the validation dataset. The data used for algorithm training and development was not eligible for evaluating the study objectives, and the data split was pre-defined prior to study start. The Liom computational procedure, LCMV1, was developed based on the data of this development cohort. The LCMV1 processes the raw signals via simple frequency transformation and filtering to ensure that the data are standardized before being analyzed by a model for estimating glucose. The development investigators (FR, AP) were blinded to the reference data used to evaluate this study's primary and secondary outcomes. A pre-defined, mutually exclusive data split was used to evaluate the primary and secondary endpoints of the study. The analysis of the validation set was performed in silico, after study completion, by an independent statistician from another company not involved in the clinical intervention or data collection (MR). All analyses were performed using the intention-to-treat dataset using the statistical package R version 4.4.0. No imputation for missing data was performed, while obviously erroneous data points based on spectral quality were excluded from the analyses.

*Point accuracy assessment*

The primary analysis was performed by visualizing both the Clinical Demo 2.0 values and the reference plasma glucose measurements over the time of intervention in individual graphs. In addition, the correlation between the two curves was assessed using rolling window time lagged cross-correlations. The determination of the accuracy of the glucose estimations against venous plasma glucose measurements was assessed by computing both the overall mean absolute relative difference (MARD) and the MARD at the participant level[2]. Parkes error grids[3] were generated for the full validation dataset and separately for each participant. We report the proportion of data points for each of the five zones A, B, C, D and E for the full validation dataset and separately per participant.

*Feature importance of the computational model*

A 64 mM glucose solution was prepared in a UV-cuvette to collect a clean Raman spectral signature of the glucose molecule using the same set up as in the clinical investigation. Raman spectra were cropped in the wave number range 227 to 3668 cm-1 and denoised.

### Suppl. Figure 1. time series of development and validation cohort

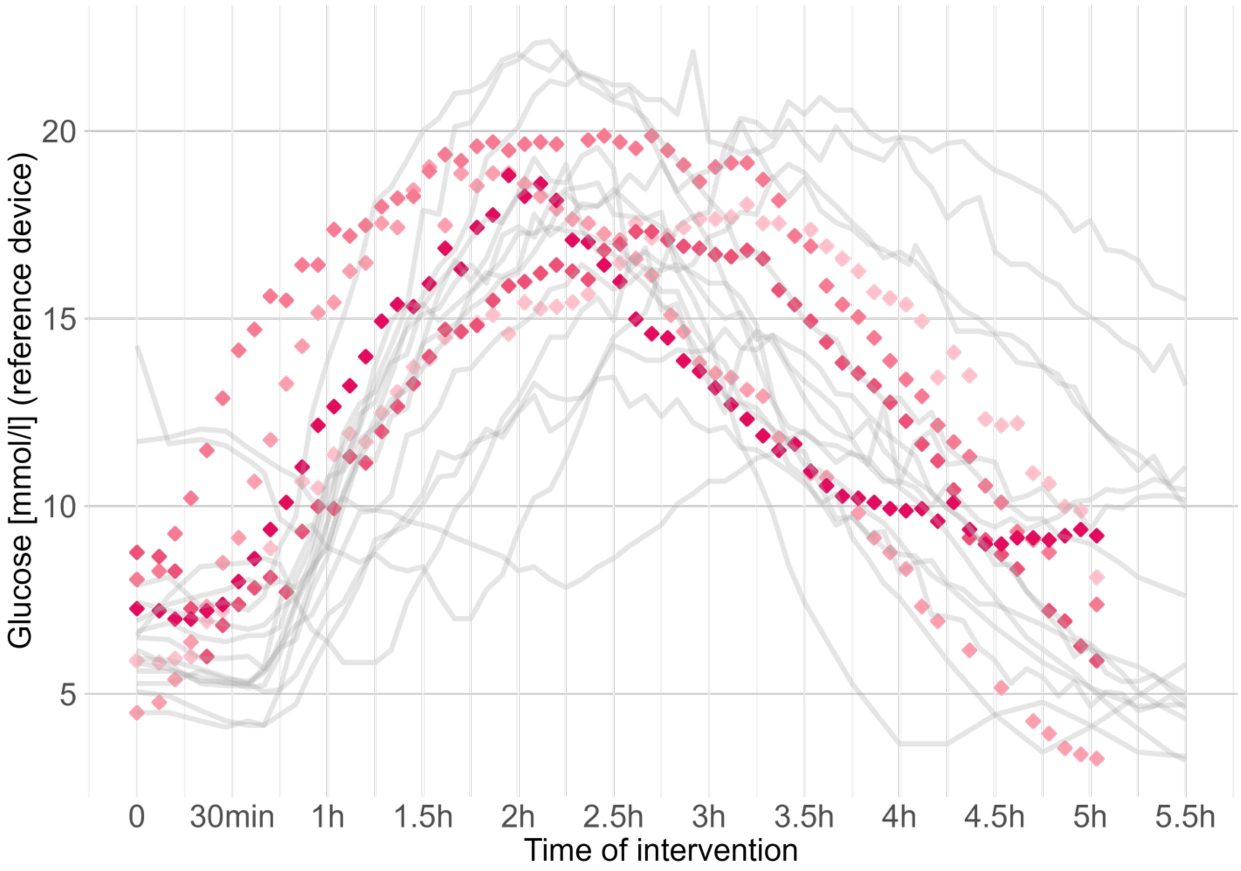


Time series in pink visualize the intervention of the validation cohort and in gray of the development cohort

### Suppl. Table 1. Eligibility criteria

Inclusion criteria:

1) Signed and dated informed consent
2) Clinical diagnosis of type 1 or type 2 diabetes for at least 1 year
3) Between 18 and 65 years old
4) Treatment with basal insulin, or with both basal insulin and oral antidiabetic drugs, multiple dosing insulin therapy (MDI), continuous subcutaneous insulin infusion (CSII) or a hybrid closed loop system
5) Body Mass Index (BMI) 18.5 to 35.0 $kg/m^2$
6) HbA1c at screening below or equal to 9.5%.
7) Sufficient venous access allowing for cannulation

Exclusion criteria:

1) Known or suspected hypersensitivity to any of the components of the investigational device
2) Presence of any injury, infection, atypical skin condition (e.g., hyperkeratosis, hyperpigmentation) of or tattoo on the wrists
3) Skin tone based on the Fitzpatrick scale V or VI
4) Application of a skin toning agent, collagen supplement, or beta-carotene supplement within the last 14 days before screening
5) Presence or history of a cardiovascular disease including stable and unstable angina pectoris, myocardial infarction, transient ischaemic attack, stroke, cardiac decompensation, clinically significant arrhythmias or clinically significant conduction disorders
6) Trial participant has an active implantable medical device, such as a pacemaker
7) Participation in any clinical trial investigating with a medical device or any medicinal product in clinical development within 30 days
8) Any history or presence of cancer except basal cell skin cancer or squamous cell skin cancer
9) Clinically relevant comorbidity, capable of constituting a risk for the trial participant when participating in the trial or of interfering with the interpretation of data as judged by the investigator
10) Presence of signs of acute illness
11) Presence of any serious systemic infectious disease during four weeks prior to the intervention

12) Presence of clinically significant abnormal screening laboratory tests or standard 12-lead electrocardiogram (ECG) after 5 minutes resting in supine position at screening
13) Significant history of alcoholism or drug abuse or consummation of more than 24.0 grams alcohol/day (for males) or 12.0 grams alcohol/day (for females)
14) Inability or unwillingness to refrain from smoking for the duration of visit 2.
15) Positive test for hepatitis Bs antigen, hepatitis C antibodies, HIV-1/2 antibodies or HIV-1 antigen.
16) Blood donation or blood loss of more than 500 mL within the last 3 months
17) Mental incapacity, unwillingness or language barriers precluding adequate understanding or co-operation
18) Pregnancy or current breastfeeding
19) Affiliation with trial site or trial personnel
20) Consideration of unsuitability for inclusion by investigator

### Suppl. Table 2. Baseline characteristics

| | Overall | Development cohort | Validation cohort |
|---|---|---|---|
| Participants | 20 | 15 | 5 |
| Women | 10 (50%) | 9 (60%) | 1 (20%) |
| Age [years] | 41 (IQR 31 to 45) | 45 (IQR 32 to 50) | 31 (IQR 27 to 41) |
| Skin color according to Fitzpatrick scale | | | |
| Type I | 0 (0%) | 0 (0%) | 0 (0%) |
| Type II | 5 (25%) | 5 (33.33%) | 0 (0%) |
| Type III | 8 (40%) | 8 (53.33%) | 0 (0%) |
| Type IV | 7 (35%) | 2 (13.33%) | 5 (100%) |
| Height [cm] | 173.5 (IQR 168 to 178) | 174 (IQR 168 to 178) | 173 (IQR 169 to 173) |
| Weight [kg] | 78 (IQR 66 to 90) | 80.5 (IQR 61 to 91) | 75.5 (IQR 75 to 89) |
| BMI [kg/m$^2$] | 25.8 (IQR 22.9 to 28.575) | 25.3 (IQR 22.8 to 28.85) | 26.5 (IQR 25.2 to 27.8) |
| HbA1c [mmol/l] | 7.3 (IQR 6.7 to 7.7) | 7.5 (IQR 7 to 7.7) | 6.7 (IQR 6.7 to 7.1) |

Numbers shown are counts (%) or medians (IQR)

### Suppl. Table 3: Procedural characteristics

| | Overall | Development cohort | Validation cohort |
|---|---|---|---|
| Subjects (n) | 20 | 15 | 5 |
| Time to hyperglycemia (min) | 52.5 (IQR 23.75 to 60) | 55 (IQR 47.5 to 60) | 25 (IQR 10 to 35) |
| Time in hyperglycemia (min) | 260 (IQR 197.5 to 274.25) | 220 (IQR 185 to 274.5) | 265 (IQR 255 to 270) |
| Time from hyperglycemia to end of intervention (min) | 22.5 (IQR 0 to 72.5) | 55 (IQR 0 to 82.5) | 15 (IQR 5 to 25) |
| Blood glucose at study start (mmol/l) | 6.5 (IQR 5.8 to 7.5) | 6.5 (IQR 5.7 to 7.2) | 7.3 (IQR 5.9 to 8) |
| Blood glucose during hyperglycemia (mmol/l) | 14.2 (IQR 13.3 to 15.5) | 14.2 (IQR 13.3 to 15.5) | 14.6 (IQR 13.8 to 14.6) |
| Blood glucose at discharge (mmol/l) | 6.6 (IQR 4.7 to 10) | 5.8 (IQR 4.7 to 10.3) | 7.4 (IQR 5.9 to 8.1) |
| Rate of change | | | |
| Low RoC | 426 (37.4%) | 329 (37.2%) | 97 (37.9%) |
| Moderate RoC | 351 (30.8%) | 272 (30.8%) | 79 (30.9%) |
| High RoC | 177 (15.5%) | 139 (15.7%) | 38 (14.8%) |
| Very high RoC | 186 (16.3%) | 144 (16.3%) | 42 (16.4%) |

Numbers shown are counts (%), medians (IQR) or minimum/maximum values
